# Using visual biofeedback to reduce step length error at fast walking speeds is feasible after stroke

**DOI:** 10.64898/2026.06.08.26355006

**Authors:** Christina K. Holl, Maryana Bonilla Yanez, James M. Finley, Andrew Hooyman, Kristan A. Leech

**Author notes:** **Corresponding author:** Kristan A. Leech, PT, DPT, PhD, Division of Biokinesiology and Physical Therapy, University of Southern California, 1540 Alcazar St., CHP 155, Los Angeles, CA 90033.

## Abstract

**Background and Purpose:** Walking after stroke is often characterized by persistent biomechanical impairments and reduced walking capacity. While visual biofeedback can improve gait mechanics and fast walking can enhance capacity, it is unclear whether individuals post-stroke can effectively use biofeedback at higher walking speeds to address both deficits simultaneously. This study examined the effects of walking speed on the ability of participants with chronic stroke to reduce step length (SL) errors using visual biofeedback.

**Methods:** Sixteen individuals with chronic stroke walked on a treadmill at slow, self-selected, and fast speeds with and without visual SL biofeedback. Absolute SL error relative to individualized targets was calculated for paretic and non-paretic limbs. Linear mixed-effects models with piecewise linear splines assessed the effects of speed, limb, and feedback condition. Post hoc comparisons were performed for significant interactions.

**Results:** At lower speeds, increasing speed reduced SL error in both limbs (p < 0.001). At higher speeds, the effects of speed were dependent on limb and condition (p < 0.001). Paretic SL error increased with speed without feedback but remained stable with feedback (p < 0.001). Non-paretic SL error decreased with speed regardless of condition. SL error was greater in the paretic limb overall (p < 0.001).

**Discussion and Conclusions:** Fast walking alone did not reduce paretic SL errors. Participants with chronic stroke can effectively use visual biofeedback to reduce paretic SL errors at higher speeds, supporting its integration into high-intensity gait training to simultaneously treat biomechanical impairments and walking capacity deficits after stroke.

## Introduction

Walking deficits are one of the most common and disabling consequences of a stroke. While most people regain some degree of walking function, many continue to demonstrate long-term biomechanical impairments and limited walking capacity. Biomechanical impairments, such as insufficient swing and increased step length asymmetry,^1,2^ are associated with important health outcomes, including an elevated fall risk.^3–5^ Similarly, limited walking capacity post-stroke, manifesting as decreased walking speed and walking distance, reduces community mobility and quality of life.^6^ Historically, physical therapy approaches have often emphasized these two domains of biomechanical impairments and walking capacity independently. While there is some evidence that biomechanical impairments are correlated with metrics of walking capacity,^7,8^ interventions to improve one domain do not necessarily result in improvements in the other.^9^ Therefore, an integrated approach that explicitly targets both the biomechanical impairments and limited walking capacity may maximize recovery for those living with stroke.

To primarily target biomechanical impairments, physical therapists often employ feedback-based training.^10^ External feedback about movement performance can be provided through visual, auditory, or haptic cues, allowing people with stroke to target specific movement errors, such as step length or paretic push-off.^11^ This training approach engages strategic and use-dependent learning processes linked to plasticity in multiple brain structures (e.g., the primary motor and dorsolateral prefrontal cortices)^12,13^. While research indicates that people post-stroke can effectively use biofeedback to modify their gait, feedback-based research is most often conducted at people’s self-selected walking speeds.^12^ In contrast, high-intensity gait training (HIGT) has emerged as the gold standard to target walking capacity limitations after stroke.^13^ HIGT involves treadmill or overground walking at fast speeds targeting high aerobic intensities.^14–16^ These intensities are thought to increase neuromotor engagement during walking and leverage transient, intensity-dependent neuroplastic changes — including increased corticospinal excitability and neurotrophin expression^17^ to enhance use-dependent motor learning in individuals post-stroke^18^ and able-bodied controls^19^. While this intervention includes strategies to address neuromuscular deficits that support walking function (e.g., stance control), it does not explicitly target specific kinematic and spatiotemporal deficits (e.g., step length asymmetry).^20^ As such, HIGT improves kinetic variables (e.g., hip and ankle power) but produces limited changes in kinematic and spatiotemporal patterns.^21^ Integrating spatiotemporal or kinematic biofeedback into HIGT could efficiently promote changes in both domains and may yield synergistic effects, given the complementary mechanisms of these interventions. However, it is unclear how well people post-stroke can use biofeedback to reduce walking-pattern impairments at fast speeds.

Using biofeedback while walking fast may be challenging for people post-stroke, as it may impose additional cognitive load to an already cognitively demanding task. Previous studies have shown that participants make more errors on a visuo-spatial cognitive task during fast walking than during self-selected walking, suggesting that fast walking is more cognitively demanding.^22^ Furthermore, biofeedback itself is cognitively demanding, as individuals must continuously process external feedback and integrate it into their ongoing motor output. Importantly, ∼60% of people have cognitive impairment after stroke,^23^ reducing the cognitive resources needed for dual tasks^24^ and has been associated with poorer performance with biofeedback during walking.^25^ Collectively, this suggests that fast walking speed may reduce one’s ability to make biofeedback-based biomechanical changes to their walking pattern after stroke.

This study aimed to evaluate the ability of individuals with chronic stroke to use visual biofeedback across different walking speeds. To do so, we examined step length (SL) error while participants with chronic stroke walked at slow, self-selected, and fast speeds, with and without visual biofeedback. We hypothesized that (1) people post-stroke would be able to reduce SL error when provided visual biofeedback regardless of speed, and (2) the magnitude of error reduction depends on walking speed, with greater error observed during faster walking.

## Methods

### Participants

We recruited individuals with chronic stroke to participate in this study. The inclusion criteria were: (1) age 18-80 years, (2) at least 6 months from stroke onset, (3) able to walk independently for at least 5 minutes, and (4) paresis confined to one side. Exclusion criteria included: (1) damage to the pons, basal ganglia, or cerebellum, (2) signs of cerebellar involvement or extrapyramidal symptoms, (3) uncontrolled hypertension, (4) inability to provide informed consent (determined by remote Montreal Cognitive Assessment score <19), and (5) orthopedic or pain conditions that affect walking ability. All participants provided informed consent prior to participation, approved by the Institutional Review Board of the University of Southern California. This study is registered on clinicaltrials.gov (NCT04411303).

### Experimental setup and protocol

Participants completed clinical measures of motor impairment (Fugl-Meyer Assessment, Lower Extremity^26,27^), balance (Berg Balance Scale^28^), endurance (6-Minute Walk Test^29^), and walking speed (10-Meter Walk Test^29^) prior to the experimental task. Then, participants walked at three different treadmill speeds for three minutes in random order: slow speed, self-selected walking speed (SSWS), and fastest safe walking speed (FSWS), with and without visual biofeedback (Figure 1A). SSWS and FSWS were determined using a custom staircase algorithm based on their overground walking speed. The slow speed was determined by multiplying the SSWS by the ratio of SSWS to FSWS. Participants first completed the Feedback Off condition, and then completed the Feedback On condition. This ordering allowed us to determine whether participants’ step lengths deviated sufficiently from the target to warrant the Feedback On condition. During the Feedback On condition, participants walked with step length biofeedback (Figure 1B).

**Figure 1.**
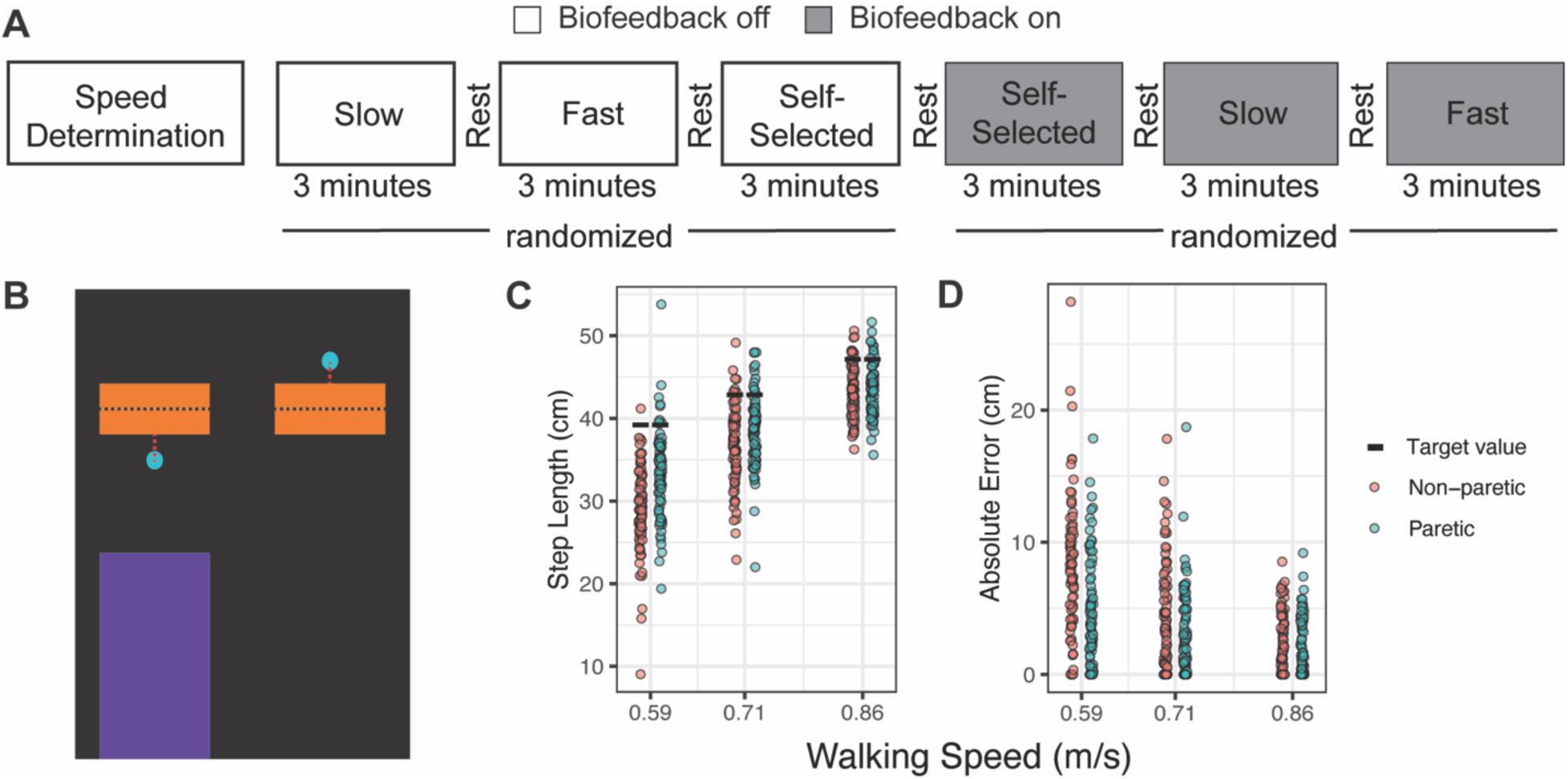
Experimental paradigm and data from representative participant. **A)** Participants walked on the treadmill at three randomized speeds, with and without biofeedback, resting as long as needed between trials. **B)** Snapshot of real-time step length biofeedback display. The purple bar represents the real-time step lengths at a given time. The blue dot provided endpoint feedback, representing the participant’s previous step length. The black dashed line was not visible to the participants and represents the individualized step length target value. The orange rectangle was visible to the participants and represents the goal zone, the width of which was set at ±5% of the target value. **C)** Step lengths for both limbs at all three speeds during the Feedback On condition for a representative participant. The black rectangle represents the participant’s step length target value. **D)** Absolute step length error across all three speeds for the same representative participant. Absolute step length error was calculated for each step as the absolute value of the distance between step length and the closest goal zone border (red vertical dashed line in panel B, not visible to participants). Abbreviations: cm, centimeter; m/s, meter/second.

Our step length biofeedback provided real-time step lengths of both limbs (Figure 1B, purple bars). It also provided step length end-point feedback, allowing participants to visualize their previous step length values (Figure 1B, blue dot). Individualized step length targets were based on the participant’s age, sex, mass, leg length, and walking speed^30^ with a ±5% tolerance goal zone (Figure 1B, orange rectangles). Participants were given the following instructions before the first biofeedback trial with a static visual of the biofeedback display: “The purple bars show how your feet are moving in real-time. The position of the blue dots represents the length of your right and left steps on the step before. The goal is to get the blue dots to land in the orange bar.” The participants were not provided with a practice session with the biofeedback before completing the biofeedback trial.

### Instrumentation and data processing

Three-dimensional motion capture data were recorded at 100 Hz (Qualisys AB, Gothenburg, Sweden). Markers were placed bilaterally on each participant’s fifth metatarsal, lateral malleoli, lateral epicondyle, greater trochanter, iliac crest, and posterior superior iliac spine. Kinematic data were gap-filled if 30 frames or less were missing and lowpass filtered with a cutoff frequency of 6 Hz. Heel strike was defined as the most anterior location of the lateral malleoli marker and toe-off as its most posterior location. Step length was defined as the fore-aft difference between the right and left lateral malleoli markers at heel strike. Subjects whose step lengths during the self-selected walking speed Feedback Off condition were ±5% of the step length target value (within the feedback goal zone) did not undergo trials with visual biofeedback and were excluded from the analysis. Additionally, trials in which subject walking speeds were less than 0.36 m/s were also excluded from the analysis, as these speeds were outside of the range of data used to train the prediction models for the subject-specific step length targets.^30^

Figure 1C displays step length data relative to individualized step length target values for a representative participant at all three walking speeds during the Feedback On condition. To quantify participants’ ability to use the biofeedback at each gait speed, we calculated absolute SL error (individual data displayed in Figure 1D), defined as the absolute distance between their actual step lengths and the nearest SL target boundary (Figure 1B, red dashed lines). To determine how speed alone affects this metric, we also calculated SL error during the Feedback Off condition, even though SL targets were not displayed. We chose to calculate absolute SL error (rather than directional error) because we were interested in the participants’ ability to adjust their SLs to achieve the provided SL target. Therefore, either over- or undershooting the SL target was considered an error. Visual inspection of individual time-series data confirmed stable performance throughout each trial; we therefore analyzed the final 82 steps per trial — the minimum across participants.

### Model Specification and Selection

We modeled absolute stepping error (AbsError) using linear mixed-effects regression in the lme4 package (Bates et al., 2015) in R (version 4.5.2; R Core Team, 2024). We iteratively increased model complexity to select the final model. We began with a model including only the main effects of Condition, Limb, and Speed. Interaction terms were then progressively added, culminating in a model including the three-way interaction among Condition, Limb, and Speed. Inclusion of a random intercept for participant and a random slope for Speed improved overall model fit, and this random-effects structure was retained for all subsequent analyses.

To characterize the functional form of the relationship between Speed and absolute stepping error, we compared several candidate representations within the same fixed- and random-effects structure. Specifically, Speed was modeled as (1) a second-degree polynomial, (2) a logarithmic transformation, and (3) a piecewise linear spline.

We then optimized the knot location for the piecewise linear spline. An initial search evaluated candidate knot positions across a range of speeds (0.50 – 1.52 m/s) to identify the value that minimized model information criteria. Given the potential for limb-specific differences in locomotor control among people with stroke, a subsequent optimization allowed the knot to vary independently for each limb. Candidate knot combinations for each limb were systematically evaluated, and the combination yielding the best model fit was selected. The final model therefore incorporated limb-specific knots, with predictors representing the trend of Speed less than the knot and the trend of Speed greater than the knot. Model fit across these alternatives was compared, and the limb-specific piecewise linear spline provided the best representation of the data, indicating that the relationship between Speed and error differed across distinct regions of the speed range.

Post-hoc comparisons were conducted based on the final spline model. Estimated marginal means of absolute stepping error were calculated separately for the regions below and above the knot by holding one segment constant while varying the other at representative values. Pairwise comparisons among Condition levels were then performed within each Limb for each speed region, with Tukey’s method used for adjustment for multiple comparisons. In addition, the rate of change of absolute stepping error with respect to Speed was estimated separately for speeds below and above the knot, and contrasts were used to test for differences in these slopes across conditions within each limb.

## Results

### Patient Characteristics

Twenty-four individuals with chronic stroke (52 ± 14 years old; 17 males, 7 females) participated in the study. Data from sixteen of these participants were included in the analyses. Two participants were unable to complete the protocol due to endurance limitations, three had baseline step lengths within the target zone; one was excluded due to technical difficulties; and two had self-selected walking speeds outside the acceptable range^30^ (see Methods Section). Demographics for the participants included in the analysis are presented in Table 1.

**Table 1.** Clinical demographics of participants included in the analyses.

| Sex | Affected side | 10m walk speed (m/s) | 6-minute Walk Distance (m) | LEFM (/34) | Berg Balance Scale (/56) | Slow Treadmill Speed (m/s) | Self-selected Treadmill Speed (m/s) | Fast Treadmill Speed (m/s) |
| --- | --- | --- | --- | --- | --- | --- | --- | --- |
| M | Right | 1.19 | 471 | 33 | 55 | 0.74 | 1.02 | 1.40 |
| F | Right | 0.69 | 289 | 25 | 53 | 0.54 | 0.69 | 0.88 |
| F | Right | 0.46 | 192 | 17 | 44 | 0.30* | 0.42 | 0.63 |
| M | Right | 0.93 | 374 | 22 | 52 | 0.63 | 0.88 | 1.22 |
| M | Left | 1.25 | 463 | 22 | 54 | 1.06 | 1.17 | 1.29 |
| M | Left | 0.50 | 184 | 19 | 44 | 0.40 | 0.50 | 0.62 |
| M | Right | 0.62 | 223 | 21 | 50 | 0.24* | 0.47 | 0.93 |
| F | Left | 1.00 | 430 | 29 | 55 | 0.79 | 0.93 | 1.09 |
| F | Left | 0.98 | 404 | 20 | 55 | 0.57 | 0.82 | 1.18 |
| M | Left | 0.67 | 412 | 24 | 48 | 0.42 | 0.72 | 1.77 |
| M | Left | 0.42 | 164 | 16 | 43 | 0.40 | 0.72 | 1.22 |
| M | Right | 1.23 | 576 | 29 | 56 | 0.85 | 1.17 | 1.62 |
| F | Right | 0.47 | 193 | 22 | 51 | 0.21* | 0.46 | 1.03 |
| F | Left | 0.48 | 303 | 23 | 47 | 0.33* | 0.53 | 0.85 |
| M | Left | 0.77 | 301 | 19 | 45 | 0.59 | 0.71 | 0.86 |
| M | Left | 1.15 | 552 | 29 | 56 | 0.76 | 1.05 | 1.45 |

Participants whose slowest speed was excluded from the analysis are denoted with an \*. Abbreviations: M, male; F, female; LEFM, Lower-Extremity Fugl-Meyer; m, meter; m/s, meter/second.

### Final Model Evaluation

A piecewise linear representation of Speed provided the best fit to the data compared to polynomial and logarithmic alternatives. Initial knot optimization identified an optimal shared knot at 0.53 m/s (AIC = 69513.63), indicating a transition point in the relationship between walking speed and absolute error. Allowing the knot to vary by limb further improved model fit. A two-dimensional grid search of candidate knot pairs revealed optimal limb-specific knots at 0.52 m/s for the non-paretic limb and 0.62 m/s for the paretic limb (Supplemental Figure 1; AIC = 69490.37). The final model, therefore, incorporated limb-specific piecewise segments of Speed, representing the trend of Speed less than the knot and the trend of Speed greater than the knot, along with their interactions with Condition and Limb, and included random intercepts and random slopes for Speed by participant.

### Effect of Visual Biofeedback and Walking Speed

Figure 2 displays the absolute step length errors for both limbs of all participants while walking at all speeds and under Feedback On and Feedback Off conditions. The model revealed significant main effects of Speed below the knot, Condition, and Limb (Table 2). The trend of Speed less than the knot was negatively associated with absolute step length error (β = −18.24, 95% CI [−21.48, −14.93], p < 0.001). Condition also showed a significant main effect, with higher error in the Feedback Off condition relative to Feedback On (β = 3.90, 95% CI [2.75, 5.05], p < 0.001). A significant main effect of Limb indicated greater error in the paretic compared to the non-paretic limb (β = 14.77, 95% CI [13.36, 16.14], p < 0.001). The main effect of the trend of Speed greater than the knot was not significant (p = 0.393).

**Figure 2.**
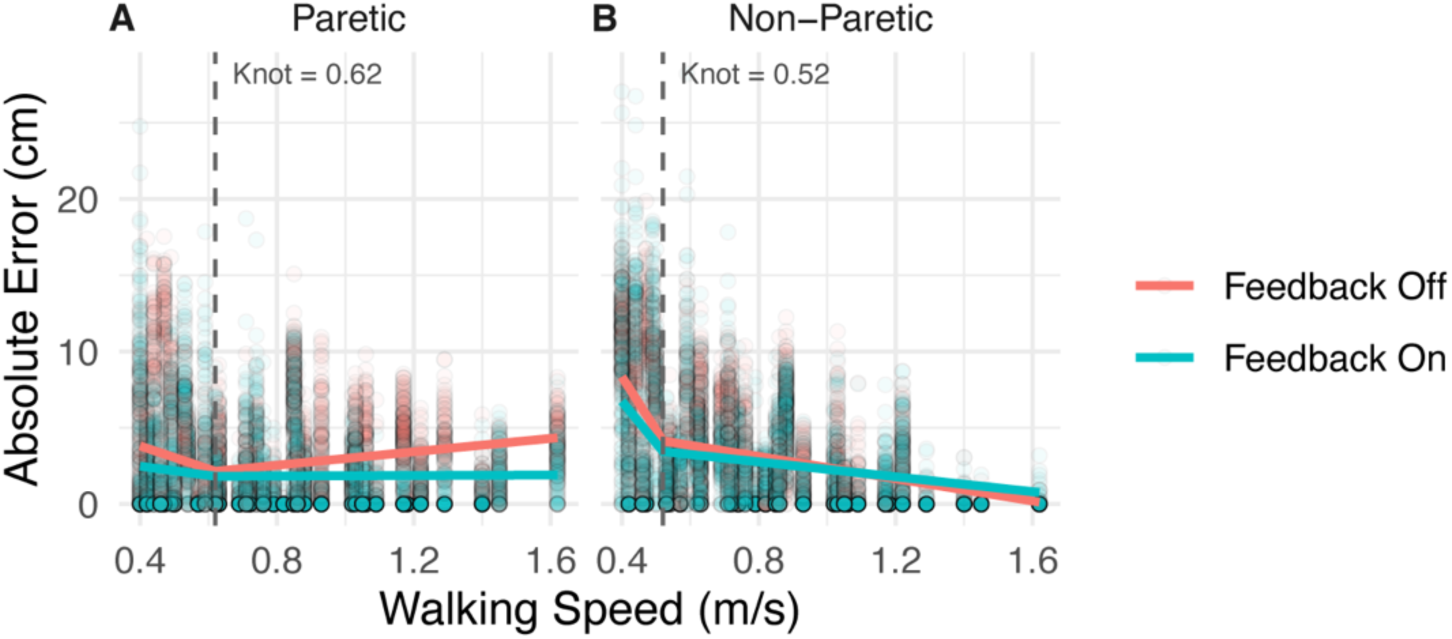
Absolute step length error values for the **A)** paretic and **B)** nonparetic limbs across speeds in both Feedback Off and Feedback On conditions. Solid color lines represent piecewise fixed-effect model fits derived from the piecewise three-way model. Vertical dashed lines signify the optimal limb-specific knot speeds. Abbreviations: cm, centimeter; m/s, meter/second.

**Table 2.**
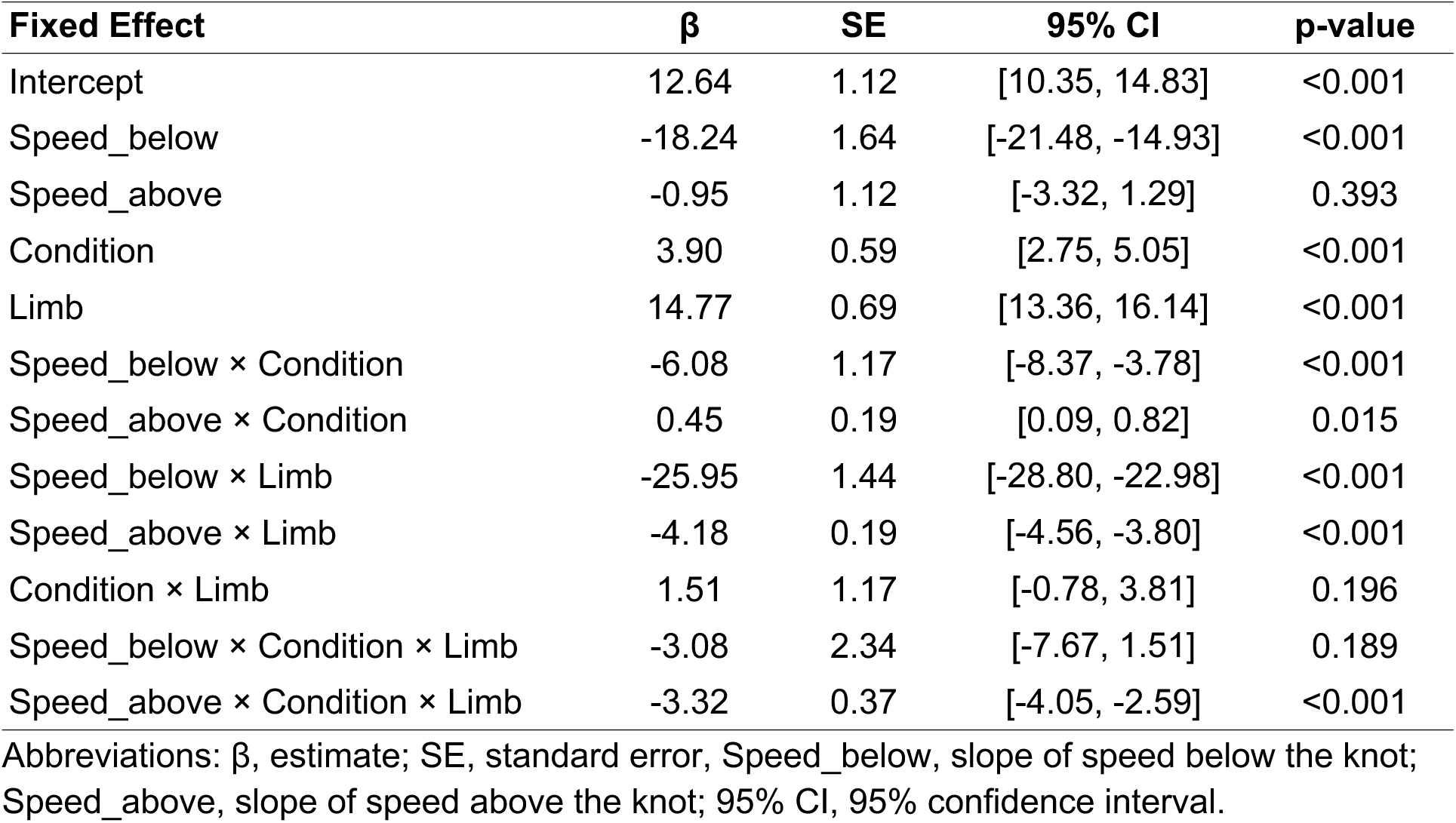
Fixed-effect estimates from the piecewise three-way model of step length error.

| **Fixed Effect** | **$\beta$** | **SE** | **95% CI** | **p-value** |
| --- | --- | --- | --- | --- |
| Intercept | 12.64 | 1.12 | [10.35, 14.83] | <0.001 |
| Speed_below | -18.24 | 1.64 | [-21.48, -14.93] | <0.001 |
| Speed_above | -0.95 | 1.12 | [-3.32, 1.29] | 0.393 |
| Condition | 3.90 | 0.59 | [2.75, 5.05] | <0.001 |
| Limb | 14.77 | 0.69 | [13.36, 16.14] | <0.001 |
| Speed_below × Condition | -6.08 | 1.17 | [-8.37, -3.78] | <0.001 |
| Speed_above × Condition | 0.45 | 0.19 | [0.09, 0.82] | 0.015 |
| Speed_below × Limb | -25.95 | 1.44 | [-28.80, -22.98] | <0.001 |
| Speed_above × Limb | -4.18 | 0.19 | [-4.56, -3.80] | <0.001 |
| Condition × Limb | 1.51 | 1.17 | [-0.78, 3.81] | 0.196 |
| Speed_below × Condition × Limb | -3.08 | 2.34 | [-7.67, 1.51] | 0.189 |
| Speed_above × Condition × Limb | -3.32 | 0.37 | [-4.05, -2.59] | <0.001 |

Abbreviations: $\beta$ , estimate; SE, standard error, Speed\_below, slope of speed below the knot; Speed\_above, slope of speed above the knot; 95% CI, 95% confidence interval.

Several interactions were significant. There was a significant interaction between the trend of Speed less than the knot and Condition (χ²(1) = 26.90, p < 0.001), as well as between the trend of Speed greater than the knot and Condition (χ²(1) = 5.91, p = 0.015). Interactions between Limb and both segments of Speed were also significant (trend less than the knot: χ²(1) = 322.57, p < 0.001; trend greater than the knot: χ²(1) = 476.32, p < 0.001). The three-way interaction between Speed less than the knot, Condition, and Limb was not significant (χ²(1) = 1.73, p = 0.189). Importantly, the three-way interaction among the trend of Speed greater than the knot, Condition, and Limb was significant (χ²(1) = 79.99, p < 0.001), indicating that differences in the speed–error relationship at higher speeds depended on both limb and condition.

To interpret the results of the three-way interaction, we performed post-hoc comparisons for speeds above the knot (Table 3), showing that mean absolute step length error differed by Condition in the paretic limb (Figure 2A) but not in the non-paretic limb (Figure 2B). For the paretic limb, mean error was higher in the Feedback Off condition (mean = 3.38, 95% CI [2.34, 4.43]) compared to Feedback On (mean = 2.01, 95% CI [0.96, 3.05]; estimate = 1.38, 95% CI [1.23, 1.53], p < 0.001). In contrast, no significant difference between conditions was observed in the non-paretic limb (Feedback Off: mean = 0.98, 95% CI [−0.05, 2.01]; Feedback On: mean = 1.13, 95% CI [0.10, 2.16]; estimate = −0.15, 95% CI [−0.44, 0.13], p = 0.290). This indicates that the participants used feedback to reduce error in the paretic limb, but not in the non-paretic limb.

**Table 3.** Post hoc comparison of means for walking speeds above the knot.

| **Limb** | **Condition** | **EMM** | **Contrast**<br>**(Off-On)** | **95% CI** | **p-value** |
| --- | --- | --- | --- | --- | --- |
| Paretic | Feedback Off | 3.38 | 1.38 | [1.23, 1.53] | < 0.0001 |
|  | Feedback On | 2.01 |  |  |  |
| Non-paretic | Feedback Off | 0.98 | -0.15 | [-0.44, 0.13] | 0.29 |
|  | Feedback On | 1.13 |  |  |  |

Abbreviations: EMM, estimated marginal mean; 95% CI, 95% confidence interval.

Analysis of the slopes above the knot further clarified this interaction (Table 4). In the non-paretic limb, absolute stepping error decreased with increasing Speed in both conditions (Feedback Off: trend = −3.65, 95% CI [−6.03, −1.27]; Feedback On: trend = −2.44, 95% CI [−4.82, −0.06]), with a significantly steeper decline in the Feedback Off condition (estimate = −1.21, 95% CI [−1.69, −0.73], p < 0.001). This suggests that in the non-paretic limb, SL errors naturally reduce with increases in gait speed (Figure 2B). In contrast, for the paretic limb, the slope was positive in the Feedback Off condition (trend = 2.20, 95% CI [−0.20, 4.59]) and near zero in the Feedback On condition (trend = 0.08, 95% CI [−2.32, 2.48]), with a significant difference between conditions (estimate = 2.11, 95% CI [1.57, 2.66], p < 0.001). This indicates that when walking without feedback, at higher speeds SL errors emerged, and participants were able to use feedback to stabilize SL error with increases in speed (Figure 2A).

**Table 4.**
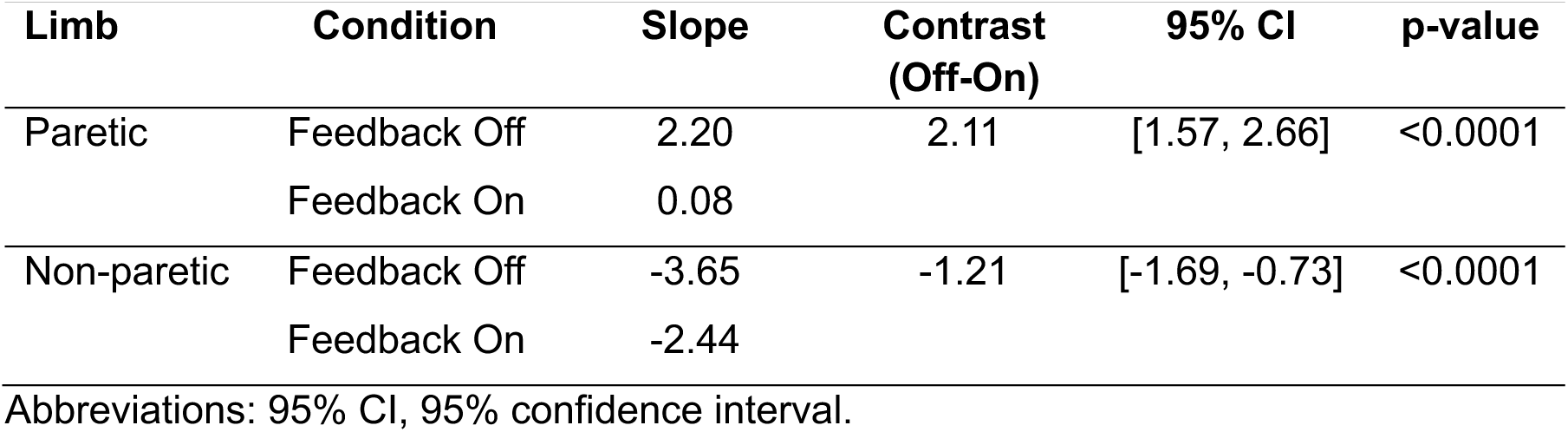
Post hoc comparison of slopes for walking speeds above the knot.

| **Limb** | **Condition** | **Slope** | **Contrast**<br>**(Off-On)** | **95% CI** | **p-value** |
| --- | --- | --- | --- | --- | --- |
| Paretic | Feedback Off | 2.20 | 2.11 | [1.57, 2.66] | <0.0001 |
|  | Feedback On | 0.08 |  |  |  |
| Non-paretic | Feedback Off | -3.65 | -1.21 | [-1.69, -0.73] | <0.0001 |
|  | Feedback On | -2.44 |  |  |  |

Abbreviations: 95% CI, 95% confidence interval.

Comparisons between limbs revealed that the slope of the trend of Speed greater than the knot differed significantly between limbs in both conditions. In the Feedback Off condition, the non-paretic limb showed a more negative slope than the paretic limb (estimate = −5.84, 95% CI [−6.37, −5.32], p < 0.001). A similar pattern was observed in the Feedback On condition (estimate = −2.52, 95% CI [−3.04, −2.00], p < 0.001), indicating fundamentally different speed–error relationships between limbs at higher walking speeds.

## Discussion

We aimed to understand how walking speed affects the use of visual biofeedback in people with chronic stroke. We found that the relationship between speed and error shifts at a specific walking speed for each limb: at slower speeds, increasing walking speed reduces SL error in both the paretic and non-paretic limbs, whereas at faster speeds the effect of speed is limb-dependent. In the non-paretic limb, participants’ SL errors continued to decrease as speed increased, regardless of the feedback condition. In contrast, paretic limb SL errors increased at faster walking speeds without biofeedback, and participants were able to use biofeedback to reduce these speed-dependent errors. This suggests that therapists may simplify the biofeedback provided to focus exclusively on the paretic limb. These results are contrary to our hypothesis and demonstrate that participants can use visual biofeedback to reduce paretic limb errors even under higher-demand walking conditions.

We expected that individuals post-stroke would have difficulty using visual biofeedback to reduce SL error at fast walking speeds due to the increased neuromotor and cognitive demands of fast walking. However, participants were able to use visual biofeedback to counteract the emergence of SL errors (with the error trend slope near zero) that occurred naturally at faster walking speeds (> 0.62 m/s) in the absence of biofeedback. This result is somewhat surprising for two main reasons. First, emerging evidence suggests that visuospatial function may be engaged during walking after stroke to a greater extent than other cognitive domains,^31^ and performance with visual biofeedback is related to visuospatial function post-stroke.^25^ Second, high-demand lower-extremity tasks increase reliance on compensatory neural control strategies after stroke,^32^ including greater muscle coactivation,^33^ which could limit the ability to modify walking patterns in response to biofeedback. Yet our results contradict this expectation. Indeed, the speed knots identified by our piecewise model align with this idea, as they indicate a speed threshold for a shift in neuromotor control of walking, particularly for the paretic limb in which we observed the emergence of paretic SL error at speeds above the knot.

Though unexpected, our results suggest that combining interventions targeting distinct domains of walking recovery is feasible. This understanding may have important clinical implications. First, post-stroke rehabilitation therapy doses remain low in the United States.^34^ Therefore, the limited number of physical therapy sessions available must be used efficiently, and a more comprehensive approach that simultaneously targets walking capacity and gait biomechanics should be considered, particularly given the potentially complementary or synergistic effects of these interventions on walking after stroke. This idea is supported by prior work demonstrating enhanced outcomes when fast walking is paired with other gait-retraining strategies after stroke. For example, combining fast walking with functional electrical stimulation has been shown to improve paretic propulsion to a greater extent than fast walking alone,^35^ and adding therapist-provided verbal feedback further enhanced motor learning during gait training.^36^ Furthermore, our work clarifies that non-paretic reductions in SL error are largely dependent on speed, allowing physical therapists to concentrate feedback strategies on the paretic limb. Collectively, these findings support the idea that high-intensity walking interventions may serve as an effective foundation upon which more impairment-specific interventions can be layered to optimize recovery. However, the present study evaluated only the immediate effects of combining fast walking with visual biofeedback. Longer-term intervention studies are needed to determine whether these acute changes translate into sustained improvements in gait biomechanics and walking capacity after stroke.

This study has a few additional limitations. First, fast walking speeds were determined by the pace participants reported they could maintain for 3 minutes, rather than at the individual’s biomechanical limits or heart rate zones, as in published protocols for high-intensity gait training.^37^ It remains unclear if participants would be able to use SL biofeedback at those speeds. Second, the Feedback Off condition always preceded Feedback On, introducing the potential for order effects. However, the speed- and limb-specific responses to biofeedback observed suggest that the effects were not solely driven by practice or task familiarization. Finally, our individualized step length targets were developed from prediction models trained on walking speeds between 0.36 and 2.23 m/s^30^ which required us to exclude data from people walking at speeds < 0.36 m/s. As a result, our findings may not generalize to people with more severe walking impairments who walk slowly.

## Conclusion

It is feasible for people with stroke to use paretic step length biofeedback to reduce errors while walking at fast speeds. We observed limb-specific speed thresholds in the relationship between walking speed and SL error. At slow speeds below the threshold, increasing gait speed naturally drove SLs closer to the individualized target values in both limbs. Above the threshold, however, SL errors increased in the paretic limb when no feedback was given. The introduction of SL feedback counteracted this trend, preventing SL error from worsening with increasing speeds. Collectively, these findings suggest that fast walking alone may be insufficient to improve paretic gait mechanics at higher speeds and support the integration of biofeedback into high-intensity gait training to simultaneously target walking capacity and biomechanical impairments after stroke.

## Data Availability Statement

Data are available from the corresponding author upon reasonable request.

