## Supplemental Figure 1 for "Using visual biofeedback to reduce step length error at fast walking speeds is feasible after stroke"

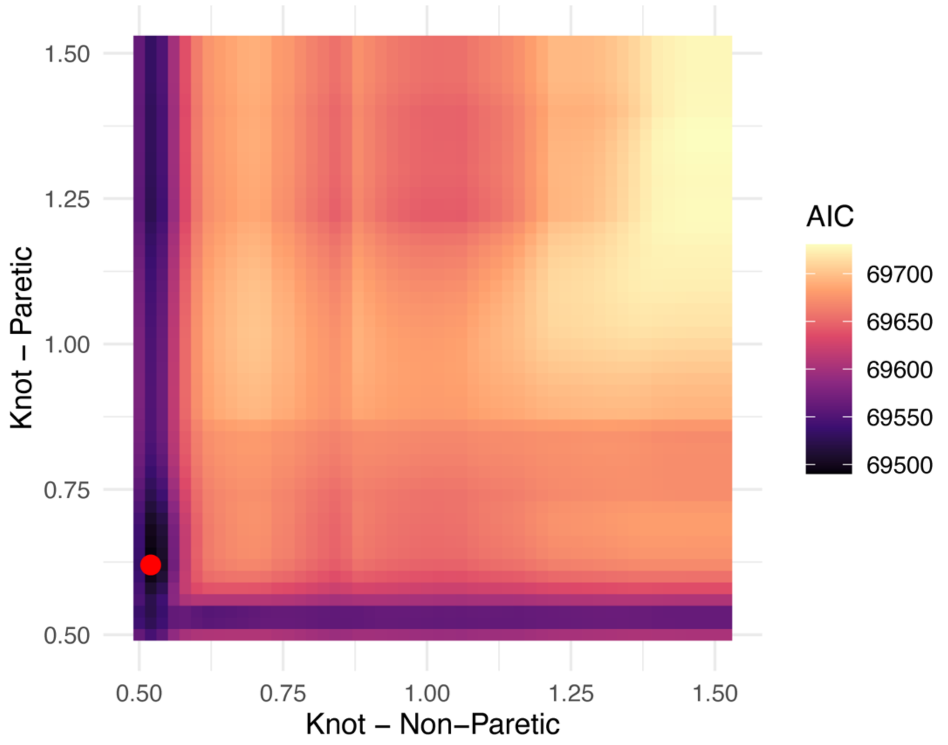


**Supplemental Figure 1.** Two-dimensional grid search heatmap of Akaike Information Criterion (AIC) across candidate knot combinations for the piecewise linear model. The x-axis and y-axis represent the limb-specific knot speeds in meters/second for non-paretic and paretic limbs, respectively. The color gradient reflects the model's AIC value for each knot pairing. Darker shades (purple/black) indicate lower AIC values (better model fit), while light shades (yellow) indicate higher AIC values. The red dot denotes the global minimum AIC for the optimal knot pairing.
